# Action observation intervention using three - dimensional movies improves the usability of hands with distal radius fractures in daily life: a nonrandomized controlled trial in women

**DOI:** 10.1101/2023.11.19.23297832

**Authors:** Kengo Usuki, Hiroaki Ueda, Toshiya Yamaguchi, Takako Suzuki, Toyohiro Hamaguchi

## Abstract

Prolonged immobilization of joints after distal radius fracture (DRF) leads to cerebral disuse-dependent plasticity (DDP) and deterioration of upper extremity function. Action observation therapy (AOT) can improve DDP. This nonrandomized controlled trial (UMIN 000039973) tested the hypothesis that AOT improves hand-use difficulties during activities of daily living in patients with DRF. Right-handed women with volar locking plate fixation for DRF were divided into AOT and Non-AOT groups for a 12-week intervention. The primary outcome was the difficulty in using the fractured hand, as examined by the Japanese version of the Patient-Related Wrist Evaluation (PRWE). Secondary outcomes were (1) range of motion (ROM) of the injured side and (2) difference between the measured and patient-estimated ROM. The survey was conducted immediately postoperatively and at 4, 8, and 12 weeks postoperatively. The AOT groups used a head-mounted display and three-dimensional video during ROM exercises, whereas the Non-AOT group used active ROM exercises alone. A generalized linear model (GLM) was used to confirm interactions and main effects by group and time period, and multiple comparisons were performed. In total, 35 patients were assigned to the AOT (n=18, median age 74 years) and Non-AOT (n=17, 70 years) groups. In the GLM, PRWE Total, PRWE Specific, and PRWE Usual scores showed interactions between groups and periods. A post-hoc test showed that the PRWE Specific (z=3.43, p=0.02) and PRWE Usual (z=7.53, p<0.01) scores w ere significantly lower in the AOT group than in the Non-AOT group at 4 weeks postoperatively, whereas PRWE Total s cores (z=3.29, p=0.04) were lower at 8 weeks postoperatively. These results suggest that AOT can improve hand-use difficulties in right-handed women after DRF surgery. AOT positively affects the motor imagery of patients with DRF and can reverse the patient’s perceived difficulty of using the fractured hand during rehabilitation.

## Introduction

Distal radius fracture (DRF) is a frequent diagnosis more common in women, occurring in 16 men and 36 women per 10,000 population [1]. DRF occurs at a younger age than hip or vertebral fractures and is known to be the first osteoporosis-related disease. Women with DRFs have a 24% higher prevalence of osteoporosis than those without. The prevalence rates of osteoporosis in people aged 50 years and over are reported to be 44% for women and 30% for men, and osteoporosis is a factor contributing to DRFs being more common in women [2] .

Sports or traffic accidents are commonly responsible for DRFs in young people, whereas falls from a standing position mainly cause DRFs in older people [3,4] . The DRF risk is 3.2 times higher in individuals who go outside at least once a day than in those who do not [5] . DRF injury is likely caused by increased activities due to roles such as housework and occupation.

For patients with DRF, goals for rehabilitation and activities of daily living (ADLs) are set one week after surgery, and instrumental ADLs such as housework are set as goals for 7-8 weeks after surgery [6] . Since patients with DRF are often highly active in home and social roles before their injury, rehabilitation is provided so that they can achieve their goals in ADL and social life as soon as possible.

Patients with markedly displaced and unstable DRFs who cannot maintain reduction undergo surgical treatment [7] . Range of motion (ROM) exercise can be performed in the early postoperative period in patients whose bones are fixed with a volar locking plate, but it takes time for patients who have undergone surgery to be able to use the fractured hand for ADLs . Bone union takes about 9 weeks in such patients [8,9], but they can use their hands for non-weight-bearing ADLs after 4 weeks. It is generally considered safe that patients exercise their wrist muscles 6 – 8 weeks postoperatively [10,11] .At 8 weeks postoperatively, approximately 80% of patients with DRF experience difficulties in using their hands for ADLs, whereas at 12 weeks, this percentage decreases to 50% [12] .

Among factors that contribute to the difficulties of patients with DRF in using their hands for ADLs are ROM l imitations [13, 14] . The ROMs of the wrist and forearm used for ADLs have been reported to be 38° for volar flexion, 40° f or dorsiflexion, 13° for internal rotation, and 53° for external rotation [13] . Patients with DRF require 4 weeks postoperatively to recover to the level of ROM used for ADLs [15] . In addition, at 4 and 8 weeks postoperatively, dorsiflexion of the wrist joint was a factor contributing to the perceived difficulty in using the operated hand for ADLs [14] . Fig 2 shows the change in wrist ROM after surgery.

Difficulty in using the affected hand in ADLs is one of the reasons why this hand is not used in ADLs. Patients with DRF underestimate their actual ROM s by − 20° volar flexion, − 21° dorsiflexion, − 31° internal rotation, and − 24° external rotation [16] . However, their ROM is limited because immobilization of the joint results in peripheral degeneration of sensory receptors [17, 18] and a decrease in superficial and positional sensation [19] . When the hands of patients with upper limb movement disorders are immobilized, the volume of the motor-sensory cortex of the brain becomes smaller approximately two weeks after injury [20], and the cerebral blood flow of the motor-sensory cortex decreases after 24 hours of joint immobilization even in healthy individuals [21] . These findings are neurophysiological evidence that immobilization of the wrist joint decreases the motor sensation of patients with DRF, and even if the ROM has improved to the angle required for ADL, patients who underestimate their ROM may refrain from using their hands or subjectively experience difficulties using them.

A decrease in hand use causes a transient decrease in local brain activity related to hand movements, which, if persistent, alters the neural structure of the brain. This phenomenon is called disuse-dependent plasticity (DDP) [22] . When a normal hand is immobilized for 2 weeks, the frequency of its use is significantly reduced, its grip strength is reduced by 36%, and its performance in an upper limb function test u sing a Purdue peg board is reduced by 21.6%. Functional magnetic resonance imaging of the brain at this time confirms DDP, in which the motor-sensory cortex of the upper limb and its surrounding network connections show reduced activity [22] .

If DDP occurs in patients with DRF and their ADLs are restricted by perceived difficulties in using their hands, training to correct this phenomenon should be considered. Rehabilitation for DRF has been performed during postoperative days 8 to 10, during which time the fracture is in the hematogenous phase, and the patient receives bracing rest and protective ROM exercises. From days 10 to 25 postoperatively, patients are in the early callus formation stage, when they receive gentle ROM exercises and perform ADLs such as eating. From postoperative days 20 to 60, the patients are in the osteoblast-proliferative phase and receive ADLs with a load of about 1 kg. From day 50 postoperatively, the patients enter the osteosclerosis phase, and at 8 weeks postoperatively, non-weight-bearing ADL exercises are performed. The postoperative period from 4 to 12 months is considered the remodeling phase, and rehabilitation is completed in about 3 months [10, 11]. Action observation therapy (AOT) is an effective means of preventing DDP in patients. AOT takes advantage of the activation of the brain cortex when observing the actions of others [23] . Rocca et al. conducted AOT of upper limb function in 42 healthy participants and reported that compared with the control group, the gray matter of the cerebrum thickened, and the upper limb function improved in the AOT group [24] . AOT in patients with DRF may prevent or reverse postoperative DDP and eliminate exacerbating factors of hand disuse. If the perceived difficulty is decreased, patients with DRF may be encouraged to use their hands for ADLs . There is no report of AOT for postoperative hand use in patients with DRF. This study tested the hypotheses that AOT in the early postoperative period of DRF can (1) prevent underestimation of ROM, (2) attenuate hand-use difficulty in ADL, and (3) improve joint ROM early in patients with DRF (Fig 3) .

## Methods

### Study design

This study was a nonrandomized controlled trial conducted between 2013. 10. 1 and 2021. 6.1. The data collection was concluded as the planned number of participants was reached.

### Ethical considerations

Written informed consent was obtained from all patients. The study was registered as a clinical trial (UMIN 000039973) and approved by the Ethics Committees of Saitama Prefectural University (25512, 2 7017) and Kitasato University Medical Center (27-57).

### Participants

The eligibility criteria for this study were women with DRF who underwent volar locking plate fixation during the study period. Exclusion criteria were (1) patients who did not wish to participate, (2) patients under 18 years of age, (3) patients with substantial soft tissue injuries in addition to fractures, (4) patients with fractures due to tumors, (5) patients with rheumatoid disorders, (6) patients with neurological diseases, and (7) patients with cognitive dysfunctions. The data collection was conducted at the Kitasato University Medical Center, during the rehabilitation.

### Experimental procedures

Study participants were assigned to two groups: one group received AOT, and the other group received the usual rehabilitation without AOT (Non-AOT). The allocation was not randomized; 10 patients were assigned to the Non-AOT group from September 2013 to August 2014 and to the AOT group from January 2016 to October 2018 for adjustment of the experimental equipment (Ghost) depending on the time of the year. Thereafter, patients were assigned to the two groups alternately in the order of their prescriptions until the sample size was met in March 2021.

### Evaluation method

In this study, the following 11 items were investigated: (1) age, (2) dominant hand (Edinburgh Handedness Inventory), (3) time from injury to start of rehabilitation (days), (4) fracture severity according to the Arbeitsgemeinschaft für Osteosynthesefragen (AO) classification, (5) body mass index, (6) injured side, (7) healthy side active ROM, (8) affected side active ROM, (9) patient’s ROM estimate for the affected side in relation to the maximum wrist ROM of the healthy side (%), (10) patient’s estimated active ROM (°), and (11) difficulty in u sing the hand for ADLs based on the PRWE, Japanese version. The survey was conducted at the time of initial rehabilitation, 4 weeks postoperatively, 8 weeks postoperatively, and 12 weeks postoperatively. The reason for this survey timing is that patients with DRF have temporary bone formation at 3-4 weeks postoperatively, whereas the fracture site stabilizes at 6 - 8 weeks postoperatively, and the orthopedic surgeon gives permission for patients to perform ADLs without restrictions. In addition, rehabilitation is completed at 12 weeks [11] . Fig 4 shows the method of measuring the difference between the patient’s estimated ROM (°) and the measured ROM (°) of the affected side, based on the maximum active ROM (100%) o n the healthy side as described previously [16] . The patient was instructed to (1) “rest the hand on the table and close your eyes” and (2) “estimate what percentage your hand moves relative to your healthy hand, assuming the healthy hand is 100%.” The reasons why patients were asked to estimate the angle percentage in comparison to the healthy side were that they ha d difficulties determining absolute angles and to prevent them from visually confirming their answers during the estimation. Test runs were not performed as underestimation might be corrected in multiple tests.

Afterward, the patient’s estimated ROM of the affected side (%) was converted into the estimated ROM (°) of the affected side using Eq.(1) :

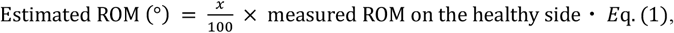

where x represents the patient’s estimated ROM (%). The difference between the patient’s estimated ROM (°) and the measured ROM (°) of the affected side was then calculated using Eq.(2):

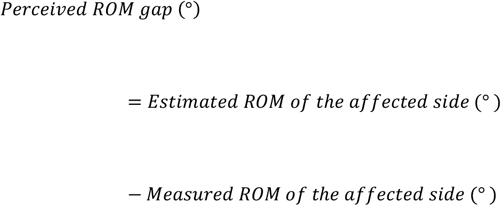

In this study, investigators were not blinded to treatment methods.

### Intervention method

The intervention was conducted for 40 minutes per session, atleast once a week, for 12 weeks after surgery. The intervention consisted of 5 minutes of evaluation, 10 minutes of passive ROM practice, another 10 minutes of active ROM practice, 10 minutes of upper limb function (ADL) practice, and 5 minutes oficing. Participants in the AOT group performed active ROM practice with the addition of AOT using a head-mounted display (HMD) and first-person three-dimensional (3D) video. AOT can consist of first-person and third-person action observations,but the first-person motion illusion is easier to produce than the third-person motion illusion, and the sense of bodily ownership of the hand on the screen is higher, as evidenced by the activity of the cerebral motor-related regions in near-infrared spectroscopy [25] . Thus, we employed an HMD-based first-person action observation system [26] .

Participants in the Non-AOT group performed active ROM exercises without AOT. Interventions were conducted by physical or occupational therapists with at least three years of experience. Fig 5 shows the posture and the equipment used during the action observation using 3D moving images. We chose 3D moving images because brain activity in response to 3D moving images is higher than that in response to 2D moving images when the participant visualizes the movement after action observation [27] .

AOT utilized an upper limb motor function learning device (Code name: Ghost, patent number 6425335, Saitama Prefectural University) [26] . Ghost comprises an HMD with personal computer software. A 3D image of another person’s hand, which had been set in advance, was presented to the patient by the HMD. The presented videos were finger flexion and extension (20 seconds),volar flexion and dorsiflexion (5 minutes), and pronation and supination (5 minutes). Patients were not instructed to perform active motor imagery during AOT.

### Sample size calculation

The sample size of this study was calculated by repeated measures analysis of variance (Cohen effect size) for group and time period, following a previous study [28], with the difference in PRWE scores as the primary outcome, after modeling the interaction between two PRWE groups and four time periods as a pre-test using a generalized linear model (GLM; f=0.25, error α=0.05, β=0.80, correlation among repeated measures=0.5, non-sphericity correction=1). The total number of cases was calculated to be 24, with 12 in each group. The number of participants prematurely terminating the study was estimated to be 20%, and the final target sample size was set to be 34 cases in total, 17 cases in each group. G*Power software (V3.1.9.6; Universität Kiel, Kiel, Germany) was used for these calculations [29] .

### Statistical analysis

The primary outcome of this study was the PRWE score as a measure of the difficulty in using hands for ADLs [30]. The secondary outcome was the difference between the estimated and measured ROM of the affected side.

The PRWE is a patient-oriented assessment of pain, difficulty in using a hand, and difficulty in using it in ADLs specific to disorders of the wrist joint. It consists of 15 questions, and the total score ranges from 0 to 100. A higher score indicates a higher degree of disability, and a score of 0 indicates no disability at all [31]. The reliability and validity of the Japanese version have been proven [32].

The Levene test was used for the test of equal variances for al l datasets, and the Shapiro– Wilk test was used to test for normality. Descriptive statistics of the characteristics of the AOT and Non-AOT groups and the primary and secondary outcomes at baseline were compared using the Mann– Whitney U test. For the primary and secondary outcomes, we used the GLM to obtain the goodness of fit of the estimated models by group and time period. When the outcome values fitted the model, multiple comparisons were tested using the Bonferroni test after checking the interaction between group and time. Age [33], body mass index [34], side of injury [35], fracture severity [36], and number of days between injury and start of rehabilitation [37] have been reported to be associated with functional outcomes,and these were used as covariates. Outliers defined as values exceeding the first quartile − quartile range × 1.5 or the third quartile + quartile range × 1.5 were excluded from statistical analyses [38] . Missing values of measurements were assigned the mean value of the group and time period [39] . Jamovi (Version 1.6.16.0) was used for data analysis [40] . The significance level was set at 0.05.

## Results

### Patient registration and analysis process

Fig 1 shows the flowchart of participant enrollment. A total of 89 patients with DRF presented during the study period, and 46 patients met the study selection criteria. A total of 11 patients were excluded from the statistical analysis because they discontinued their participation and were unable to attend the outpatient clinic. Therefore, data from 35 patients (18 in the AOT group and 17 in the Non-AOT group) were statistically analyzed. In all patients, a fall caused the injury. An outlier test was performed before further statistical analyses, and the excluded data were: 14 in the AOT group and 17 in the Non-AOT group for the measured ROM of the injured side (4 types, 4 time periods), 9 in the AOT group and 13 in the Non-AOT group for the difference between estimated and measured ROMs (4 types, 4 time periods), and 9 in the Non-AOT group for the PRWE score (4 types, 4 time periods). Table 1 shows the characteristics of the AOT and Non-AOT groups. The characteristics did not differ between the AOT and Non-AOT groups.

**Table 1.**
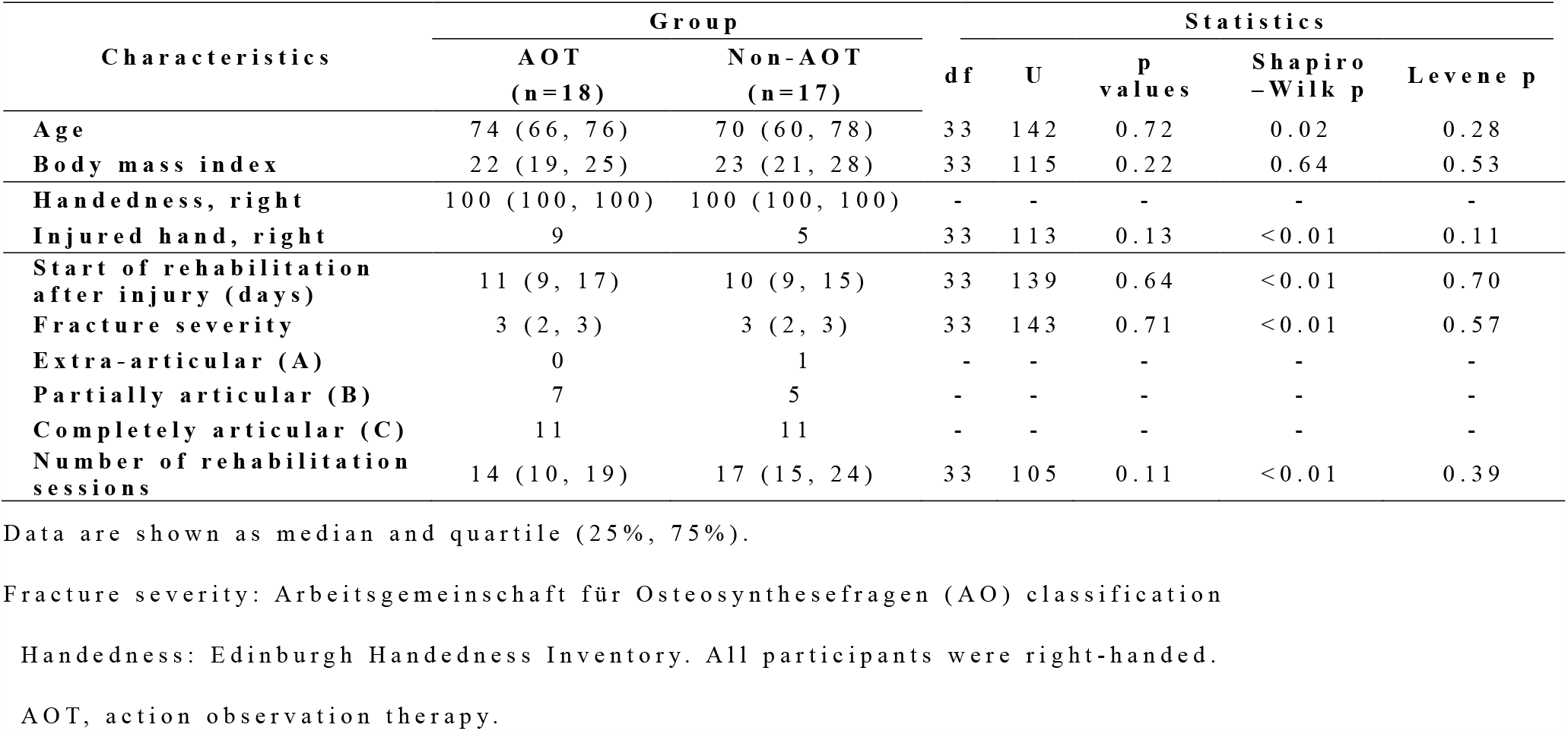
Characteristics of the study participants.

| Characteristics | Group |  | Statistics |  |  |  |  |
| --- | --- | --- | --- | --- | --- | --- | --- |
|  | AOT<br>(n=18) | Non-AOT<br>(n=17) | df | U | p<br>values | Shapiro<br>-Wilk p | Levene p |
| Age | 74 (66, 76) | 70 (60, 78) | 33 | 142 | 0.72 | 0.02 | 0.28 |
| Body mass index | 22 (19, 25) | 23 (21, 28) | 33 | 115 | 0.22 | 0.64 | 0.53 |
| Handedness, right | 100 (100, 100) | 100 (100, 100) | - | - | - | - | - |
| Injured hand, right | 9 | 5 | 33 | 113 | 0.13 | <0.01 | 0.11 |
| Start of rehabilitation<br>after injury (days) | 11 (9, 17) | 10 (9, 15) | 33 | 139 | 0.64 | <0.01 | 0.70 |
| Fracture severity | 3 (2, 3) | 3 (2, 3) | 33 | 143 | 0.71 | <0.01 | 0.57 |
| Extra-articular (A) | 0 | 1 | - | - | - | - | - |
| Partially articular (B) | 7 | 5 | - | - | - | - | - |
| Completely articular (C) | 11 | 11 | - | - | - | - | - |
| Number of rehabilitation<br>sessions | 14 (10, 19) | 17 (15, 24) | 33 | 105 | 0.11 | <0.01 | 0.39 |

Data are shown as median and quartile (25%, 75%).
Fracture severity: Arbeitsgemeinschaft für Osteosynthesefragen (AO) classification
Handedness: Edinburgh Handedness Inventory. All participants were right-handed.
AOT, action observation therapy.

**Fig 1.**
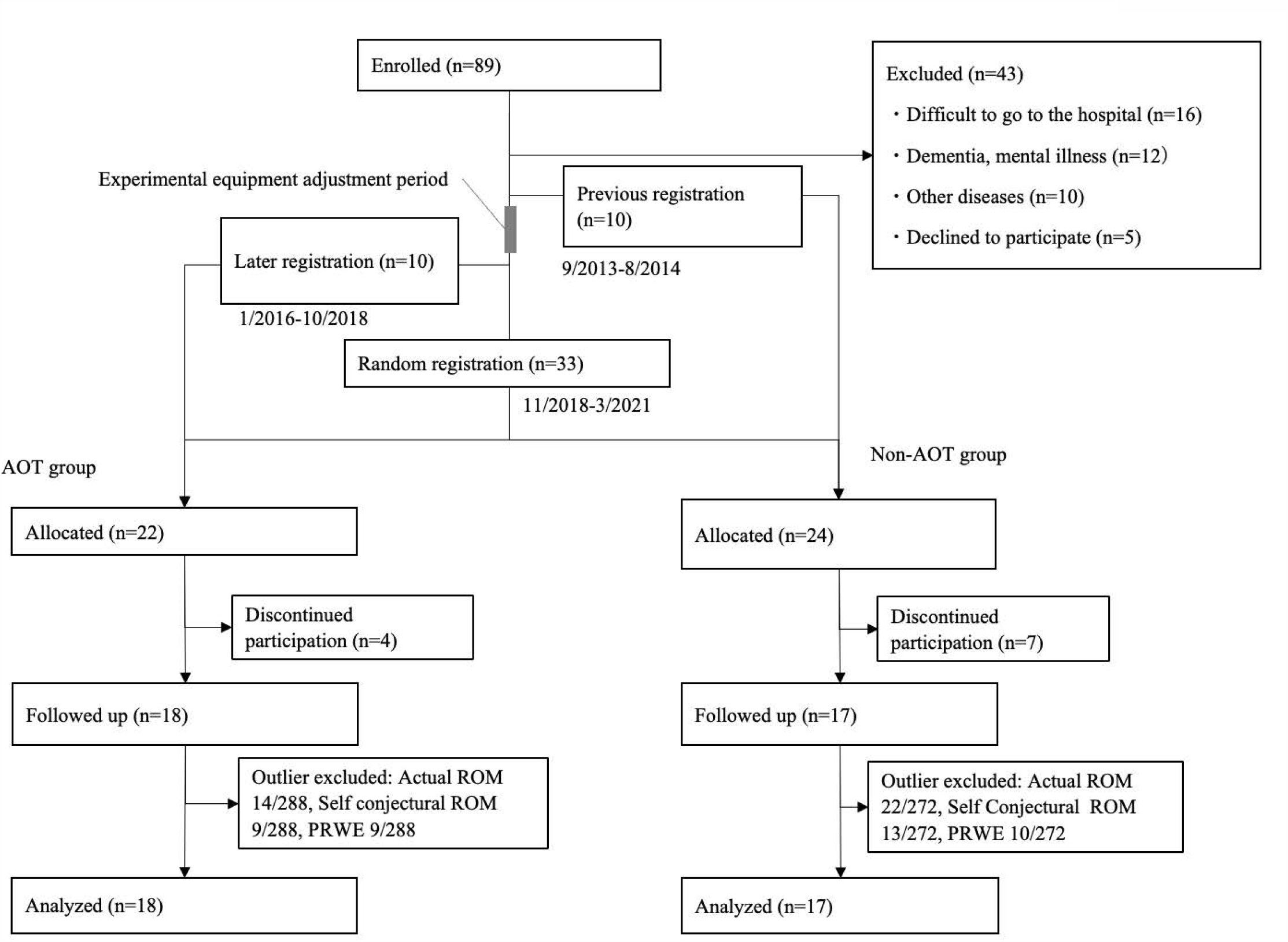
Flowchart of participant enrollment. During the study period, 89 postoperative DRF patients were admitted to the hospital, and 46 of them met the eligibility criteria. Of these, 11 patients were excluded because they were no longer able to attend the outpatient clinic. Finally, 35 patients were followed up, and their data were statistically analyzed. AOT, action observation therapy; DRF, distal radius fracture; HMD, head-mounted display; PRWE, Patient-Related Wrist Evaluation; ROM, range of motion.

**Fig 2.**
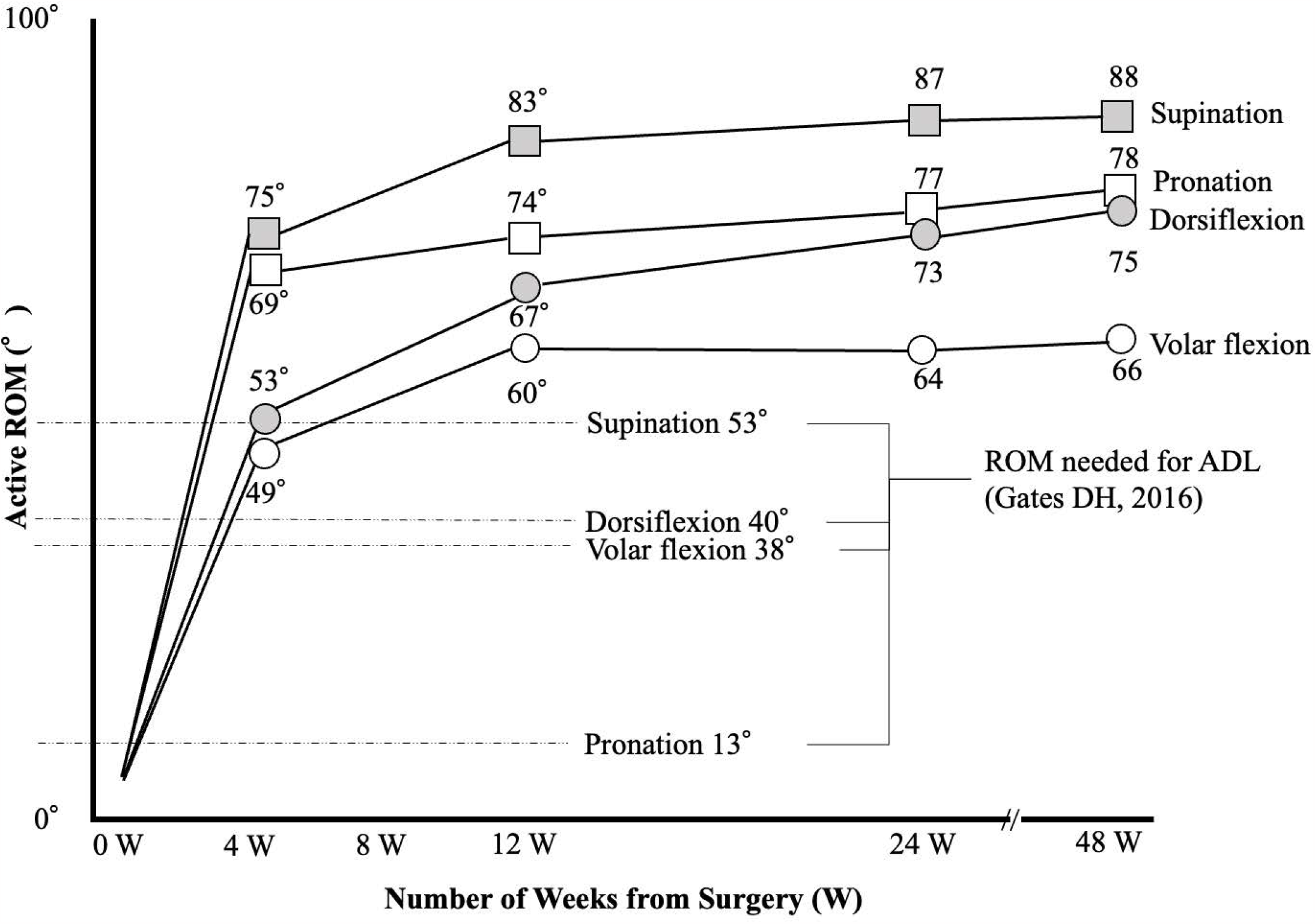
Range of motion of the wrist joint required for activities of daily living. The minimal angle for each movement is indicated by a dashed line. According to prior literature (Yang Z, 2018), patients with distal radius fractures achieve that range of motion on average within one month after surgery. ADL, activities of daily living; ROM, range of motion; W, week.

**Fig 3.**
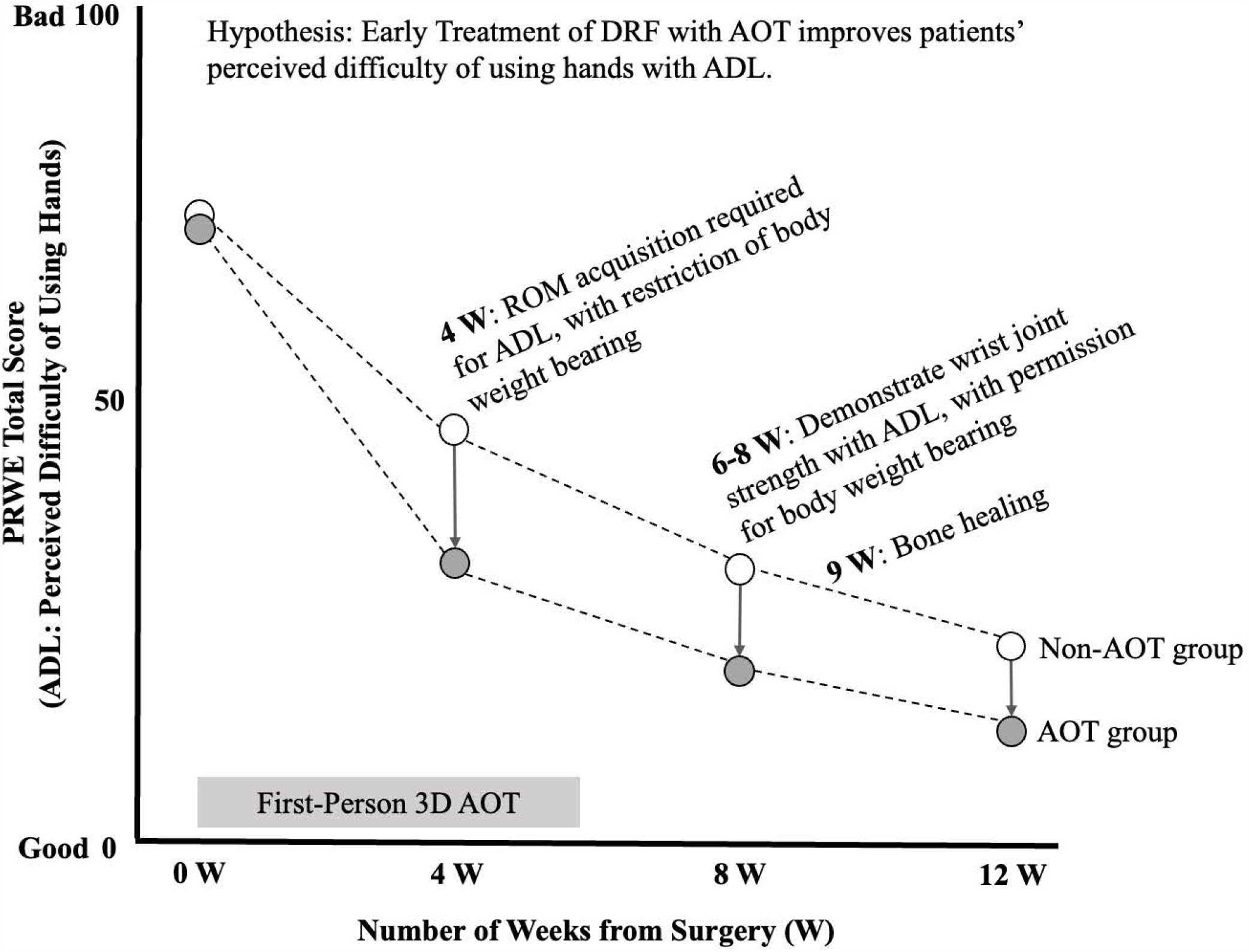
Research hypotheses. AOT in the early postoperative period improves hand use difficulty in patients with DRF from 4 weeks after surgery when patients are allowed to use their hands. In DRF, bone fusion occurs at 9 weeks postoperatively, and patients are allowed to exercise their muscles for ADLs at 6 - 8 weeks postoperatively. PRWE scores are shown on the vertical axis. The horizontal axis indicates the time after surgery. ADL, activities of daily living; AOT, action observation therapy; DRF, distal radius fracture; PRWE, Patient-Related Wrist Evaluation; ROM, range of motion; W, week.

**Fig 4.**
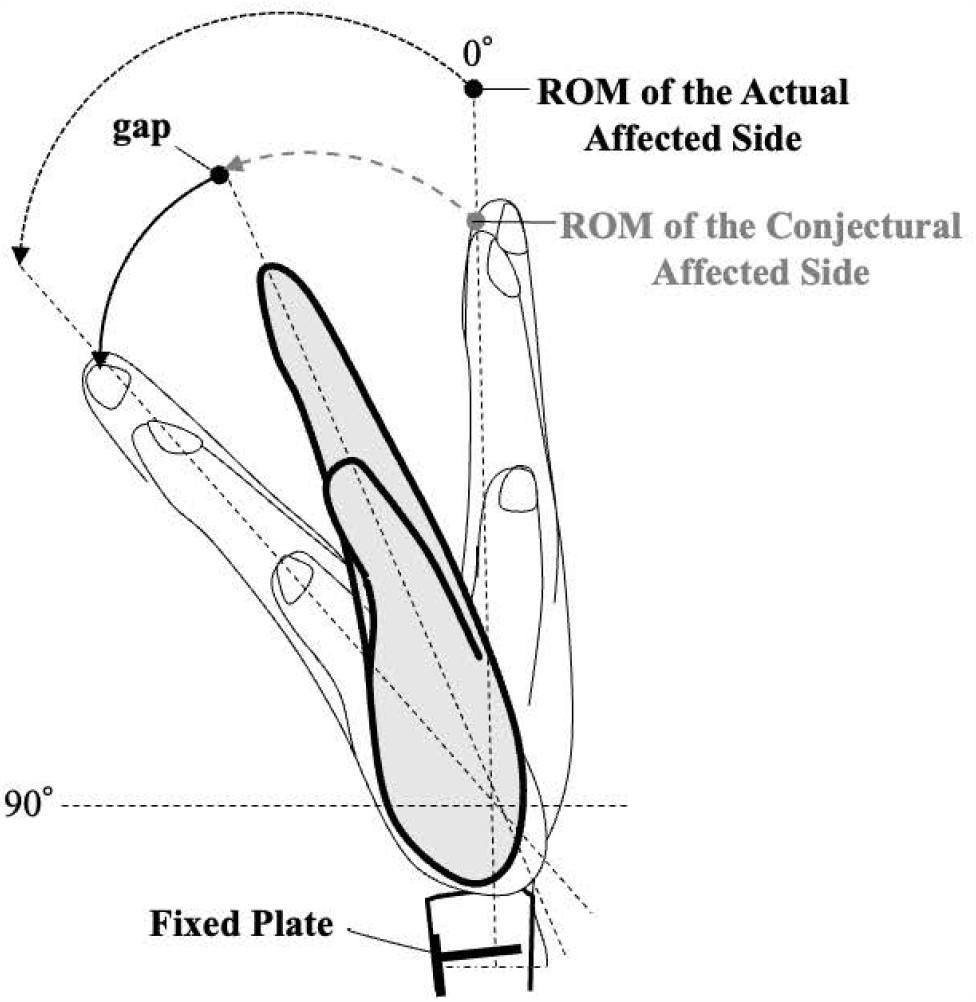
Difference between estimated and measured ROM of the affected side in wrist flexion. Patients were asked to report the estimated ROM (%) of the affected side when the healthy side was set as 100%. The patient’s estimated ROM (%) was converted into the estimated ROM (°) of the affected side based on the measured ROM (°) of the healthy side. The difference (°) was obtained by subtracting the estimated ROM from the measured ROM of the affected side. ROM, range of motion.

**Fig 5.**
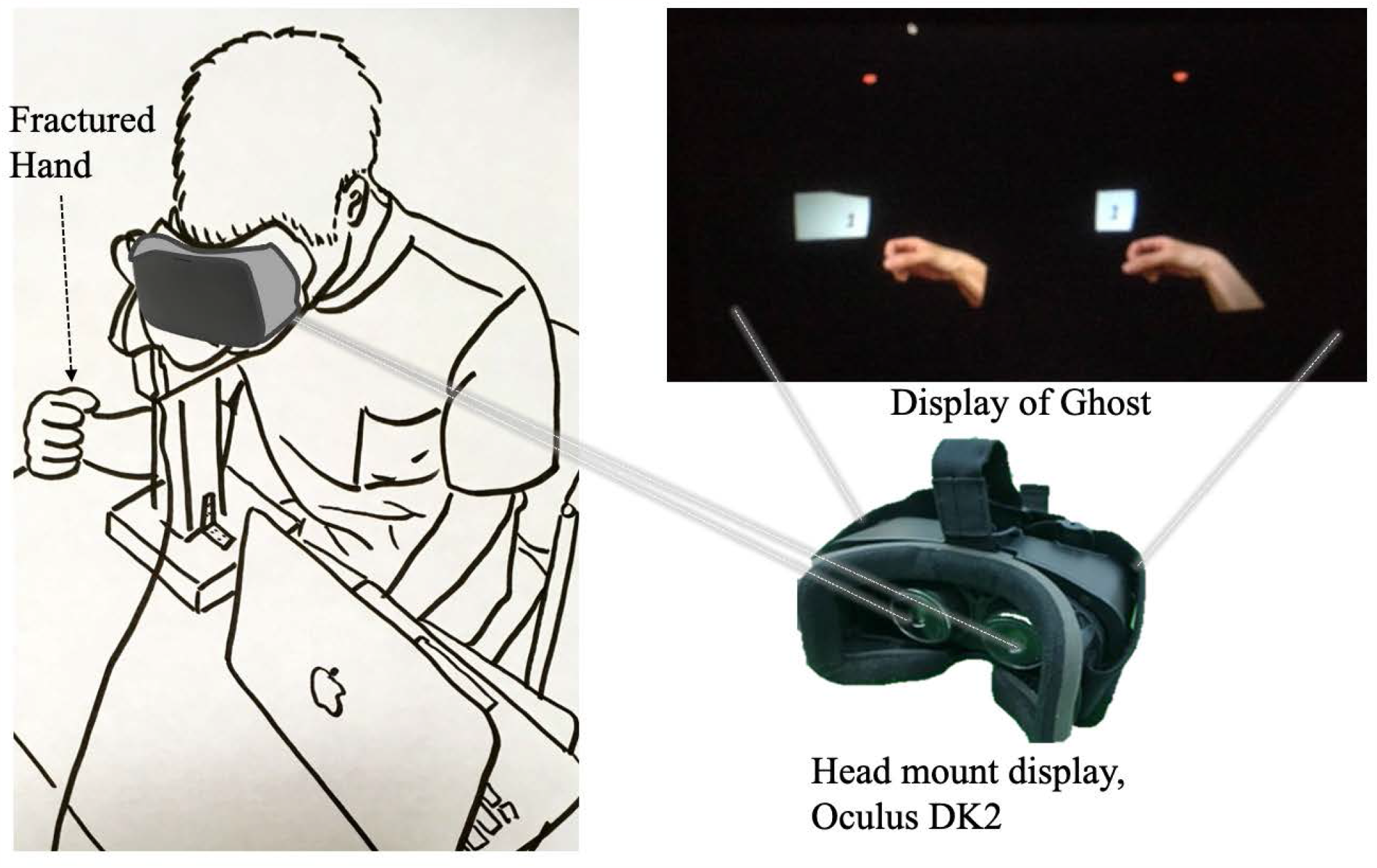
Patient’s posture during the Ghost intervention. The patient was fitted with an HMD, and AOT was performed using first-person 3D images. During the AOT, the patient practiced ROM with the fractured hand on the table in accordance with the Ghost video. AOT, action observation therapy; HMD, head-mounted display; ROM, range of motion.

### PRWE and ROM baseline data

Table 2 shows the baseline values of the measured ROM on the healthy side, measured ROM of the affected side, estimated ROM of the affected side, difference between estimated and measured ROM of the affected side, and PRWE score in the AOT and Non-AOT groups . The values for the measured ROM on the healthy side, measured ROM of the affected side, estimated ROM of the affected side, difference between estimated ROM and measured ROM of the affected side, and PRWE scores were not significantly different between the AOT and Non-AOT groups at baseline. The normality of the baseline data was evaluated using the Shapiro– W ilk test; PRWE Pain (W=0.97, p=0.41) and PRWE Total (W=0.96, p=0.19) scores had a normal distribution, whereas PRWE Specific (W=0.75, p<0.01) and PRWE Usual (W=0.90, p<0.01) scores did not. Therefore, the tests for PRWE, ROM, and difference between estimated and measured ROMs were conducted after the GLM fit was confirmed.

**Table 2.**
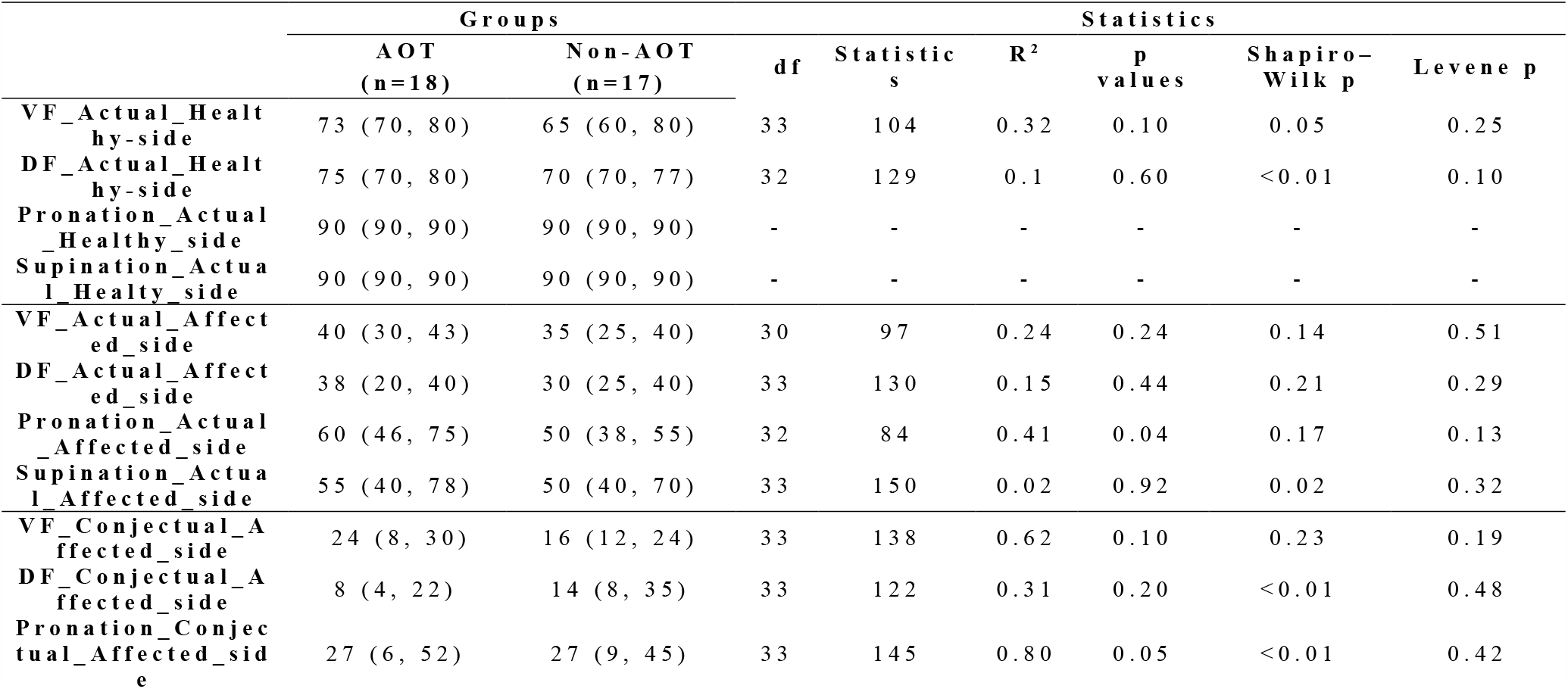
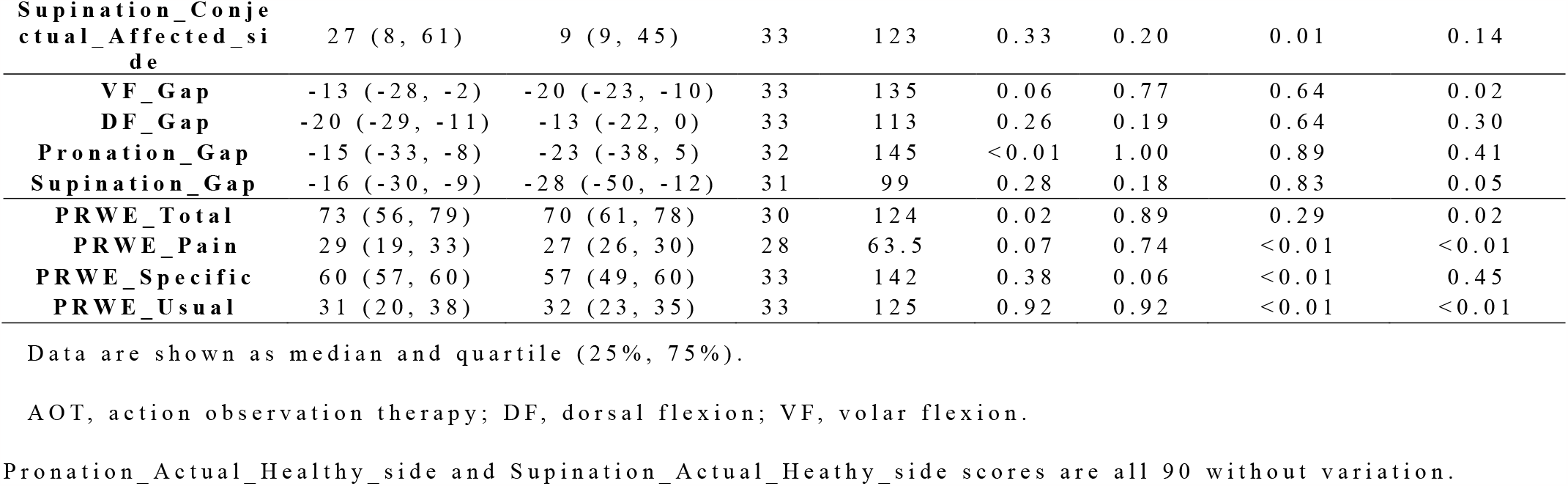
Comparisons of gaps between conjectural and actual range of motions among groups at baseline.

### Variation of PRWE and ROM by group and time period: GLM fit

Table 3 shows the changes in measured ROM, estimated ROM, difference between estimated and measured ROMs,and PRWE scores of the AOT and Non-AOT groups over time. Table 4 shows the comparative results of the ROM of the affected side, difference between estimated and measured ROMs,and PRWE scores of the AOT and Non-AOT groups by GLM. Fig 6 shows the comparison of temporal PRWE changes between the AOT and Non-AOT groups.

**Table 3.** Group comparisons for actual ROM, conjectural ROM, gap between actual and conjectural ROM, and PRWE score.

|  | Baseline |  | Postop. 4W |  | Postop. 8W |  | Postop. 12W |  |
| --- | --- | --- | --- | --- | --- | --- | --- | --- |
|  | AOT | Non-AOT | AOT | Non-AOT | AOT | Non-AOT | AOT | Non-AOT |
|  | (n=18) | (n=17) | (n=18) | (n=17) | (n=18) | (n=17) | (n=18) | (n=17) |
| **VF_Actual_Affected_side** | 40 (30, 43) | 35 (25, 40) | 55 (50, 60) | 48 (40, 55) | 60 (56, 65) | 50 (45, 60) | 63 (56, 70) | 60 (50, 60) |
| **DF_Actual_Affected_side** | 38 (20, 40) | 30 (25, 40) | 58 (54, 60) | 55 (48, 60) | 65 (60, 70) | 60 (50, 60) | 65 (60, 70) | 60 (60, 60) |
| **Pronation_Actual_Affected_side** | 60 (46, 75) | 50 (38, 55) | 90 (73, 90) | 80 (75, 90) | 90 (90, 90) | 80 (70, 90) | 90 (90, 90) | 90 (70, 90) |
| **Supination_Actual_Affected_side** | 55 (40, 78) | 50 (40, 70) | 90 (85, 90) | 80 (60, 90) | 90 (90, 90) | 90 (86, 90) | 90 (90, 90) | 90 (90, 90) |
| **VF_Conjectural_Affected_side** | 24 (8, 30) | 16 (12, 24) | 48 (35, 59) | 48 (36, 52) | 54 (48, 56) | 49 (42, 56) | 56 (48, 67) | 56 (48, 65) |
| **DF_Conjectural_Affected_side** | 8 (4, 22) | 14 (8, 35) | 43 (29, 56) | 38 (24, 49) | 51 (41, 72) | 48 (42, 56) | 56 (44, 63) | 56 (49, 63) |
| **Pronation_Conjectural_Affected_side** | 27 (6, 52) | 27 (9, 45) | 77 (45, 89) | 63 (45, 72) | 81 (72, 90) | 72 (45, 81) | 86 (81, 90) | 86 (72, 90) |
| **Supination_Conjectural_Affected_side** | 27 (8, 61) | 9 (9, 45) | 63 (48, 84) | 68 (54, 72) | 81 (72, 90) | 81 (72, 90) | 81 (72, 90) | 81 (63, 90) |
| **VF_Gap** | -13 (-28, -2) | -20 (-23, -10) | -8 (-13, -2) | -4 (-6, 2) | -10 (-17, -2) | 0 (-8, 5) | -2 (-14, 5) | 1 (-6, 10) |
| **DF_Gap** | -20 (-29, -11) | -13 (-22, 0) | -16 (-24, -5) | -11 (-21, -9) | -10 (-24, 0) | -11 (-17, 0) | -13 (-19, 2) | -2 (-4, 0) |
| **Pronation_Gap** | -15 (-33, -8) | -23 (-38, 5) | -9 (-27, 0) | -16 (-18, -9) | 0 (-18, 0) | -17 (-26, -9) | 0 (-9, 0) | 0 (-9, 0) |
| **Supination_Gap** | -16 (-30, -9) | -28 (-50, -12) | -18 (-31, 0) | -8 (-20, 0) | -9 (-17, 0) | -9 (-18, 0) | -9 (-17, 0) | 0 (-12, 0) |
| **PRWE_Total** | 73 (56,<br>79) | 70 (61,<br>78) | 38 (27,<br>45) | 50 (37,<br>68) | 20 (11,<br>28) | 37 (28,<br>54) | 14 (6,<br>23) | 18 (8,<br>37) |
| **PRWE_Pain** | 29 (19,<br>33) | 27 (26,<br>30) | 17 (11,<br>21) | 24 (13,<br>28) | 9 (6,<br>15) | 21 (9,<br>26) | 7 (3,<br>11) | 7 (4,<br>18) |
| **PRWE_Specific** | 60 (57,<br>60) | 57 (49,<br>60) | 28 (21,<br>32) | 40 (32,<br>52) | 14 (7,<br>20) | 26 (22,<br>32) | 9 (1,<br>16) | 11 (4,<br>27) |
| **PRWE_Usual** | 31 (20,<br>38) | 32 (23,<br>35) | 10 (7,<br>14) | 24 (23,<br>26) | 4 (3,<br>8) | 10 (6,<br>23) | 4 (1,<br>6) | 1 (0,<br>10) |

Data are shown as median and quartile (25%, 75%).
AOT, action observation therapy; DF, dorsal flexion; postop., postoperative; PRWE, Patient-Related Wrist
Evaluation; ROM, range of motion; VF, volar flexion; W, week.

**Table 4.** Comparisons of gaps between conjectural and actual range of motions between the study groups.

|  | Model information |  | Model result |  | Effect size | 95% CI |  | Statistics |  |
| --- | --- | --- | --- | --- | --- | --- | --- | --- | --- |
|  | R<sup>2</sup> | AIC | χ<sup>2</sup> | p | Exp(B) | Lower | Upper | z | p |
| PRWE_Total | 0.66 | 1164 | 8.10 | 0.04 | <0.01 | -31.46 | -2.84 | -2.34 | 0.02 |
| PRWE_Pain | 0.45 | 979 | 2.38 | 0.50 | 0.08 | -10.74 | 5.60 | -6.2 | 0.54 |
| PRWE_Specific | 0.73 | 1038 | 16.79 | <0.01 | <0.01 | -29.88 | -9.12 | -3.68 | <0.01 |
| PRWE_Usual | 0.68 | 931 | 14.63 | <0.01 | <0.01 | -20.00 | -5.91 | -3.61 | <0.01 |
| VF_Actual_Affected_side | 0.58 | 998 | 2.35 | 0.49 | 293.19 | -3.59 | 14.95 | 1.20 | 0.23 |
| DF_Actual_Affected_side | 0.71 | 975 | 2.12 | 0.55 | 3.05 | -7.37 | 9.60 | 0.26 | 0.80 |
| Pronation_Actual_Affected_side | 0.53 | 1110 | 2.35 | 0.50 | <0.01 | -20.99 | 4.93 | -1.21 | 0.23 |
| Supination_Actual_Affected_side | 0.54 | 1082 | 4.02 | 0.26 | 64193.96 | -2.39 | 24.53 | 1.61 | 0.11 |
| VF_Gap | 0.27 | 1058 | 6.82 | 0.08 | <0.01 | -23.40 | 0.06 | -1.95 | 0.05 |
| DF_Gap | 0.13 | 1100 | 1.58 | 0.67 | 429.34 | -6.23 | 18.36 | 0.97 | 0.34 |
| Pronation_Gap | 0.17 | 1153 | 1.85 | 0.60 | 4.833 | -15.77 | 18.91 | 1.25 | 0.21 |
| Supination_Gap | 0.23 | 1183 | 7.76 | 0.04 | <0.01 | -38.40 | -4.58 | -2.49 | 0.02 |

Data are shown as average and quartile (25%, 75%).
AIC, Akaike information criterion; CI, confidence interval; DF, dorsal flexion; VF, volar flexion.
Distribution: Gaussian

**Fig 6.**
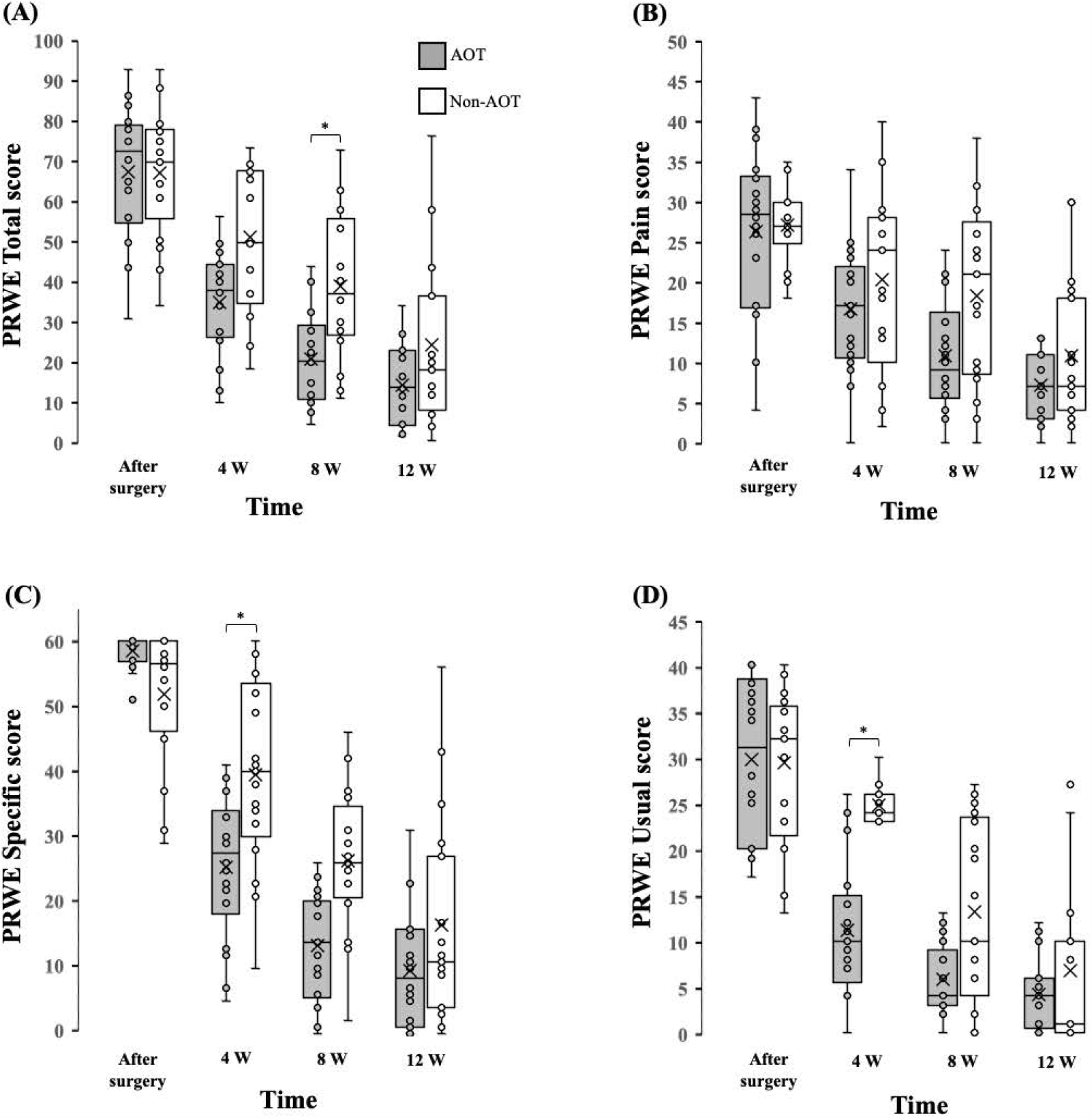
Comparison of PRWE scores between the AOT and Non-AOT groups. (A, B) PRWE Total and PRWE Pain scores did not differ between the AOT and Non-AOT groups. (C)The PRWE Specific score was significantly lower in the AOT group than in the Non-AOT group at 8 weeks postoperatively. (D) PRWE Usual scores did not differ between the AOT and Non-AOT groups. *Bonferroni, post-hoc test after GLM fitting, p<0.05. AOT, action observation therapy; GLM, generalized linear model; PRWE, Patient-Related Wrist Evaluation; W, week.

### Differences in PRWE scores between the AOT and Non-AOT groups

PRWE Total (R^2^ =0.66, Akaike information criterion [AIC]=1164, χ^2^ =8.10, p=0.04), PRWE Specific (R^2^ =0.73, AIC=1038, χ^2^ =16.79, p<0.01), and PRWE Usual (R^2^ =0.68, AIC=931, χ^2^ =14.63, p<0.01) showed a significant interaction between group and time. There was no interaction between group and time for PRWE Pain (R^2^ =0.45, AIC=979, χ^2^ =2.38, p=0.50).

Bonferroni post-hoc tests showed that the PRWE Total score was 20 [11, 28] (median [25%, 75% quartile]) in the AOT group and 37 [28, 54] in the Non-AOT group at 8 weeks (Z=3.29, p=0.04), the PRWE Specific score was 28 [21, 32] (median [25%, 75% quartile]) in the AOT group and 40 [32, 52] in the Non-AOT group at 4 weeks (Z=3.42, p=0.02), and the PR WE Usual score was 20 [7, 14] (median [25%, 75% quartile]) in the AOT group and 24 [23, 26] in the Non - AOT group at 4 weeks (Z=3.42, p=0.02).

### Differences in measured ROM of the affected side between the AOT and Non-AOT groups

The active ROM s of the affected side differed for volar flexion (R^2^ = 0. 58,AIC= 998,χ^2^ = 2.35,p=0. 49), dorsiflexion (R^2^ =0. 71,AIC= 975,χ^2^ = 2.12,p=0. 4 9), pronation (R^2^ =0. 53,AIC=1110,χ^2^ = 2.35,p=0. 50), and supination (R^2^ =0. 54,AIC=118 2,χ^2^ = 4.02,p=0. 2 6), without significant difference between the AOT and Non-AOT groups .

### Difference between estimated ROM and measured ROM in the AOT and Non-AOT groups

Regarding the differences between the estimated and measured ROM of the affected side, supination (R^2^ =0.23, AIC=1183, χ^2^ =7.76, p=0.04) showed a significant interaction between group and time.

For volar flexion (R^2^ = 0.2 7,AIC= 1 058,χ^2^ = 6.82,p=0. 08), dorsal flexion (R^2^ =0. 13,AIC=1100,χ^2^ = 1.58,p=0. 67), and pronation (R^2^ =0. 17,AIC= 1 153,χ^2^ = 1.85,p=0.60),no difference was observed between the AOT and Non-AOT groups.

The Bonferroni post-hoc test showed that supination was not significantly different at any group and time.

## Discussion

The results of this study suggest that when right-handed women with DRF receive AOT using first-person 3D video immediately after surgery, their perceived difficulty in using the affected hand in daily life and their sense of difficulty in daily life are significantly improved at 4 weeks postoperatively compared with patients without AOT. Since AOT can maintain the sense of use of the hand by affecting the motor imagery of patients with DRF, it is expected to promote the use of this hand in ADL during postoperative rehabilitation after DRF surgery.

The results of neurological studies in which AOT improved the perceived difficulty of using the injured hand in ADLs suggest that AOT increases neural excitation from the primary motor cortex to muscles via corticospinal tracts [41] and prevents cortical DDP [24] even without actual movement. Rocca et al. performed AOT for upper limb function in 42 healthy participants and reported that compared with the control group, the gray matter of the cerebrum thickened and the upper limb function improved in the AOT group [24] . Fadiga et al. compared the motor evoked potentials of the extensor digitorum communis, flexor digitorum superficialis, first dorsal interosseus, and opponens pollicis in 12 healthy individuals when they observed someone holding and reaching for an object, as well as when they observed a stationary object. Motor evoked potentials did not increase during the observation of a stationary object but during the observation of a hand releasing or reaching for an object [41] . These studies support the neurophysiological point of view that AOT is effective in preventing DDP even in the early postoperative period when the pain is strong and the patient is unable to use the affected hand due to bone fragility limiting its use in ADLs .

Opie et al. restrained the dominant index finger of healthy participant s for 8 hours and found a decrease in cortical excitability and a decrease in fine hand movements, while motor skill learning was similar to that of participants without 8 hours of restraint. The improvement in cortical excitability and fine hand movements was greater following immobilization [42] . van de Ruit et al. also investigated cortical excitability after visual motor learning and reported that the area of transcranial magnetic stimulation-induced upper limb movements increased by 18-36% after motor l earning, indicating that brain plasticity had occurred [43] . DDP is a phenomenon in which cortical excitability is reduced by fixation or disuse [20, 21], whereas use-dependent plasticity is a phenomenon in which cortical excitability is increased by upper limb function practice and motor learning u sing vision, resulting in plasticity. We assume that the Ghost-based action observation and active ROM exercises of the hand joints were effective through use-dependent plasticity preventing DDP.

In the present study, the patients’ difficulties in using the affected hand for ADLs improved at 4 weeks postoperatively, when they began to actively use the hand. At 8 weeks after DRF surgery, the patient is allowed to use the affected hand with weight bearing, suggesting that AOT intervention effectively lowers the perceived difficulty in u sing the hand for ADLs and social activities from this time onward. Dilek et al. reported improvements in pain, ROM, and ADL (Michigan Hand Questionnaire, Disability Arm Shoulder and Hand) at 8 weeks after DRF surgery following motor imagery tasks in addition to the usual rehabilitation program [44] . Thus, AOT is effective as an early postoperative intervention without exercise in the early postoperative period after DRF surgery.

MacDermid et al. reported that median PRWE Total scores after DRF surgery changed over time, with a score of 43.5 at 8 weeks postoperatively [12] . The AOT group in the present study had a median PRWE Total score of 21 at 8 weeks, suggesting an improvement compared with the results of the previous study. However, the ROM underestimation was not reduced by AOT indicating that AOT affects the perception of hand use but does not affect the actual ROM and its estimated value.

One of the reasons why AOT did not significantly improve ROM underestimation in patients with DRF is the influence of surgical intervention. I t has been reported that the flexor digitorum longus tendon, which is elongated during dorsiflexion, is impaired after volar locking plate surgery [45] . Regarding forearm rotation, the square iliacus muscle is located at the site of the surgical wound, and the ROM may have been influenced by the surgical manipulation.

DDP in patients with DRF may also be influenced by changes in peripheral sensation after a fracture. Karagiannopoulos et al. reported that the hands of patients with DRF showed decreased superficial sensation and proprioception compared with healthy hands [19] . Michinaka et al. reported a decreased number of sensory receptors with typical morphology and an increased number of those with atypical morphology in the anterior cruciate ligament after 6 weeks of knee joint immobilization in rabbits. Following 24 weeks of subsequent mobilization, these numbers were not significantly different from their baseline values although the number of atypical nerve endings remained high [18] . Nencini and Ivanusic found that the periosteum and bone marrow contain many receptors for mechanical stimuli, receptors for chemical stimuli, and receptors for pain and heat, which are involved in somatosensory perception [46] . If types and quantities of sensory receptors are similarly altered in patients with DRF, it can be inferred that sensory information from the periphery is mistakenly processed in the brain, possibly causing DDP.

Recovery after DRF is divided into the inflammatory phase, the bone fusion repair phase, and the remodeling phase. Marsell and Einhorn state that the remodeling phase begins at 3 to 4 weeks in humans and animal models and may take up to several years [47] . During the 12-week period investigated in the present study, the peripheral nerve endings and the periosteum of tissues related to somatosensory perception were not completely repaired but remained impaired, suggesting that AOT had little effect on the underestimation. AOT did not influence the measured ROM of the affected side. Brehmer and Husband compared 81 postoperative patients with volar locking plate fixations divided into two groups starting passive ROM and muscle strengthening exercises at 2 or 6 weeks postoperatively. Compared with the group that started at 6 weeks postoperatively, the group that started at 2 weeks improved in ROM, muscle strength, and ADLs (Disability Arm Shoulder and Hand score) at 8 weeks [48] . In the present study, no difference between the AOT and Non-AOT groups was found regarding ROM and muscle strength with AOT alone, suggesting the lack of an intervention effect on the actual measured ROM in the affected hand.

### Study l imitations

This was a nonrandomized controlled trial, and information bias cannot be completely eliminated. The intervention was successful in older, right-handed women; whether it is equally effective in younger women or men, or left-handed patients, needs to be examined separately. Pain and grip strength are factors affecting ADLs after DRF based on the results of a study in patients immobilized for 6 weeks with K - wire from weeks 6 to 24 [49] . Although previous studies reported differences in sensation and grip strength, the present study did not take patient-specific weight load into consideration when collecting data. According to the guidelines, maximal grip strength is not recommended until 4 weeks. In the pre-experiment, the sensation was not different between fractured and non fractured hands, indicating that sensation is not a problem. It is known that motor imagery abilities are decreased in older individuals [50, 51] . It is possible that the elderly may not be able to adapt to the changes in motor function caused by a DRF due to their reduced motor imagery ability and that the approach in this study may have facilitated improvement. Moreover, it remains unclear whether the AOT-induced improvement in ADLs with the affected hand was due to motor imagery effects or the actual movements . The difference between the effects of 2D and 3D images is also not clarified in this study. Furthermore, differences in AOT effects in relation to cognitive function, mental function, fracture severity, and the presence or absence of periosteal damage were not clarified and should be examined in future studies.

## Conclusions

In addition to the usual rehabilitation, AOT with first-person 3D moving images in postoperative women with DRF can improve the perceived difficulty in using hands for ADL and daily life at 4 and 8 weeks postoperatively. As this study was not a randomized controlled trial, the efficacy of this intervention must be verified in follow-up studies, including whether it is effective regardless of age and gender.

## Data Availability

The data underlying the results presented in the study are available from URL https://osf.io/869kr/.

## Acknowledgments

The authors would like to thank Associate Professor Toshiyuki Ishioka and Professor Kenichi Tanaka of the Graduate School of Saitama Prefectural University for their guidance in the preparation of this paper. We thank the staff of the Department of Orthopedics, Kitasato University Medical Center, the Department of Physical Therapy and Occupational Therapy, Rehabilitation Center, and Toda Central Rehabilitation Clinic for their cooperation in this study. This work was supported by JSPS KAKENHI Grant Number 26750226 and 22K21243.

## Data and study protocol Availability Statement

The data that support the findings of this study are openly available in raw_data_usuki at [DOI 10.17605/OSF.IO/869KR]. Study protocol are available in supporting information file.

